# Multi-omics Analysis Reveals Prognostic Biomarker Candidates for Calcific Uremic Arteriolopathy Patients Treated with Stem Cells

**DOI:** 10.1101/2024.12.11.24309074

**Authors:** Xiaoxue Ye, Shijiu Lu, Lianju Qin, Yaoting Sun, Jing Zhang, Ming Zeng, Jingjing Wu, Jiaying Hu, Feng Chen, Kang Liu, Yanggang Yuan, Chun Ouyang, Hongqing Cui, Lu Li, Ling Zhang, Yongwu Yu, Weigang Ge, Haibin Ren, Li Zhang, Jingfeng Zhu, Youjia Yu, Cui Li, Zhonglan Su, Dan Luo, Shaowen Tang, Xinfang Tang, Meihua Liao, Guicun Fang, Anning Bian, Fan Li, Xiying Mao, Yugui Cui, Chunyan Jiang, Xiang Ma, Song Ning, Zhanhui Gao, Baiqiao Zhao, Daoxu Wu, Cuiping Liu, Xiuqin Wang, Ningxia Liang, Changying Xing, Jiayin Liu, Tiannan Guo, Yi Zhu, Ningning Wang

## Abstract

Calciphylaxis, also known as calcific uremic arteriolopathy (CUA), is an orphan disease without proven therapies, we rescued it with human amnion-derived mesenchymal stem cells (hAMSCs). In a discovery cohort of 10 uremic patients and 3 CUA patients, plasma proteomic analysis showed core differentially expressed proteins (DEPs) Thrombospondin 1 (THBS1) and Latent transforming growth factor (TGF)-β binding protein 1 (LTBP1) decreased significantly after 3 days of hAMSC treatment. Single-cell transcriptome sequencing of peripheral blood mononuclear cells (PBMCs) indicated megakaryocytes were the source of THBS1 in CUA patient. Same as the discovery cohort, plasma THBS1 and TGF-β1 levels were increased in seven CUA patients compared to the uremic group (n=20), as measured by enzyme-linked immunosorbent assay (ELISA) in the validation cohort. They can be inhibited after hAMSC treatment and increased as the frequency of therapy decreased. THBS1 and its receptor, CD47, were increased in the CUA skin. THBS1 and TGF-β1 are biomarker candidates for calciphylaxis.

---

Calciphylaxis, also known as calcific uremic arteriolopathy (CUA), mainly occurs in chronic kidney disease (CKD) patients and is a rare, progressive, ischemic, and painful orphan disease (ORPHA:280062). In the early stages, CUA manifests as plaques, nodules, reticulated or gravid purpura, and petechiae. As the disease advances, it can progress to necrotic ulcers with black eschar.(1) In 1961, Selye et al. proposed the concept of “calciphylaxis” for the first time.(2) In uremic rats, skin tissue showed calcium deposits after exposure to sensitizing agents, but no skin ischemic necrosis was observed.(2) However, human calciphylaxis is not an inducible hypersensitivity reaction disease, and currently, there is no clinically relevant animal model for calciphylaxis. The pathological features include calcification, fibrointimal hyperplasia, and microthrombosis of dermal and subcutaneous arteries and arterioles.(3) Skin biopsy is the golden standard for definitive diagnosis, but it is invasive. In patients undergoing maintenance hemodialysis (HD), the incidence of calciphylaxis was 3.49 per 1,000 patient-years,(1) while the 1-year mortality rate is as high as 80% due to sepsis.(4)

To date, there is no approved therapeutic guideline for calciphylaxis, and a multidisciplinary combination is required.(5) Therapeutic options such as sodium thiosulfate (STS),(4) bisphosphonates,(5) and anticoagulants have been reported to be effective.(6) As a reductant, STS may reduce calcium deposition by forming water-soluble complexes with calcium(7). In some studies, STS has shown certain therapeutic effects, but side effects such as nausea, vomiting, metabolic acidosis, and volume overload need to be considered(5, 8). Furthermore, recent meta-analyses have shown that STS(9) and bisphosphonates(10) are not significantly associated with improvements in skin lesions and survival in patients with CUA compared to other commonly utilized treatments. SNF472, a specific calcification inhibitor that prevents the formation of hydroxyapatite crystals, has been shown to reduce coronary artery calcification in dialysis patients, but its efficacy in CUA remains under investigation.(11) Cinacalcet is used to treat secondary hyperparathyroidism (SHPT), potentially reducing the risk of calciphylaxis by lowering parathyroid hormone (PTH) levels and decreasing calcium-phosphate deposition, although whether it can alter the disease course remains unclear.(12) For infectious calciphylaxis, it is necessary to optimize the dialysis regimen and perform surgical debridement.(5) Patients often experience severe pain, which requires multimodal analgesia.(6) In some non-healing wounds, hyperbaric oxygen therapy may be considered as a second-line treatment option.(5, 13) In certain critical situations, amputation may be required to control infection or reduce necrotic tissue(14), but this is an extreme treatment option and is typically used as a last resort.

Currently, studies on calciphylaxis are primarily a series of case reports or retrospective studies(1, 13, 15) and exploration regarding biomarkers and novel therapeutic measures is still lacking. Stem cell-based regenerative therapies exhibit promising potential in restoring injured tissues.(16, 17) Our team has reported, for the first time, the successful rescue of CUA patients with human amnion-derived mesenchymal stem cells (hAMSCs) through intravenous and local treatment.(18) We hope to identify potential non-invasive biomarkers that can be effectively integrated into specific clinical scenarios. This investigation contributes to enhancing the diagnosis and management of challenging diseases, ultimately contributing to the optimization of therapeutic strategies, particularly those involving hAMSCs.

## EXPERIMENTAL PROCEDURES

### Ethics approval and consent to participate

All participants provided written informed consent, in accordance with the guidelines outlined in the Declaration of Helsinki.(19) This study was approved by the ethics committee of the First Affiliated Hospital of Nanjing Medical University in China (2018-QT-001, 2020-QT-01, 2020-QT-09, 2020-12-02). All procedures carried out in studies involving human participants were conducted in accordance with the ethical standards set out by their respective institutions.

### Patients and samples

We recruited ten patients diagnosed with CUA: one from the Department of Nephrology, Beijing Chuiyangliu Hospital; one from the Department of Orthopaedics, Yingkou Yanghe Hospital; and the remaining eight patients from the Department of Nephrology of the First Affiliated Hospital of Nanjing Medical University, China, between September 2018 and May 2024. Three patients with CUA were in the discovery cohort, and seven patients with CUA were in the verification cohort. We also included 30 age- and sex-matched uremic patients, whose time for renal replacement therapy was matched with CUA from the First Affiliated Hospital of Nanjing Medical University, Nanjing BenQ Medical Center, Lianyungang Oriental Hospital and the First People’s Hospital of Lianyungang. Of the uremic participants, 10 underwent plasma proteomics analysis as the discovery cohort, and another 20 uremic patients underwent plasma ELISA verification measurements for DEPs as the verification cohort. We also enrolled an age-matched healthy control for single-cell RNA-seq analysis of PBMCs.

Diagnostic criteria for CUA patients include the presence of skin lesions with severe pain, palpable subcutaneous swellings that may develop into solid hard nodules, necrotizing ulcers, or dry gangrene. Typically, these lesions appear as stellate malodorous ulcers with black eschars. (4) Skin pathology in CUA is characterized by calcification, microthrombosis, and fibrointimal hyperplasia of small dermal and subcutaneous arteries and arterioles.(1, 3) The inclusion criteria for uremic patients included: (1) aged 18 to 70 years; (2) chronic kidney disease and the etiology of uremia is “chronic glomerulonephritis.” The exclusion criteria for uremic patients included: (1) history of malignant tumor, mental illness, severe cardiovascular disease, shock, abnormal liver function, and/or secondary renal disease; (2) infection; (3) thromboembolism, coagulation function abnormalities, stenosis of arteriovenous fistula, diabetic foot and other related diseases; (4) currently using anticoagulant medication; (5) pregnant or lactating women; (6) participation in other clinical studies within the past 3 months; (7) refusal to sign informed consent.

The diagnosis and inclusion of patients with CUA and patients with uremic were done by three senior clinicians. CUA cases and uremic controls were frequency matched according to age and sex for biomarker screening and validation.

CUA patients 1-6 and 8-9, from the First Affiliated Hospital of Nanjing Medical University, were unresponsive to conventional calciphylaxis therapies.(8) They underwent salvage therapy using hAMSCs. CUA patient 7 and CUA patient 10, both of whom did not receive hAMSC therapy, were from Beijing Chuiyangliu Hospital and Yingkou Yanghe Hospital, respectively. When evaluating the efficacy of hAMSC treatment, patient skin wound healing was the primary outcome and anticoagulant medication was not adjusted. Follow-up began at the time when the patients received hAMSC treatment. Regular clinical follow-up, including outpatient visits, telephone and video calls, as well as online consultation, were scheduled for them, with a maximum period of 20 months.

Data collection was completed through the electronic medical record system, laboratory tests, blood, and skin pathology analyses. The analyses of the samples were tested in the same category and conducted in the same laboratory to guarantee comparability of measurements.

### Production of hAMSCs and rescue for CUA patients

hAMSCs were prepared in the State Key Laboratory of Reproductive Medicine, Center of Stem Cell Research and Clinical Practice of our hospital, a Good Manufacturing Practice (GMP)-compliant laboratory, according to national guidelines.(20) hAMSCs were administered intravenously to the CUA patients at a dosage of 1.0 × 10^6^ cells/kg body weight, combined with local intramuscular injection along the wound edge (2.0 × 10^4^ cells/cm^2^).(18, 21, 22)

### Collection and measurement of blood samples

Venous whole blood samples were drawn in the morning after overnight fasting from calciphylaxis patients and uremic patients. All blood samples were stored at -80°C. Routine blood tests were performed using an LH-750 Hematology Analyzer (Beckman Coulter, Fullerton, CA, USA). Serum biochemical indices were measured using an automatic biochemical analyzer (AU5400; Olympus Corporation, Tokyo, Japan). Serum intact parathyroid hormone (iPTH) levels were measured with a second-generation iPTH assay kit (UniCel DxI800 Access Immunoassay System; Beckman Coulter, Fullerton, CA, USA). Hypersensitive C-reactive protein (hs-CRP) levels were measured using an Image 800 (Beckman Coulter, Fullerton, CA, USA).

### Static and dynamic plasma proteomic sequencing and analysis of the discovery cohort

We conducted static plasma proteomic analyses on CUA patients, including patient 1, patient 2, patient 3, as well as patients with uremia (n=10). Two samples were taken from CUA patient 1, while one sample each was taken from CUA patients 2 and 3. The plasma samples of 10 uremic patients were mixed into the following groups: uremia 1, 2, and 3; uremia 4, 7, and 8; uremia 5 and 6; and uremia 9 and 10. This resulted in a total of four uremic samples for proteomic analysis. Additionally, dynamic analyses were performed on patient 1 with CUA before treatment with hAMSCs, at 3 days, 2 weeks, 1 month, and 15 months after treatment. After removing high-abundance proteins and concentrating, lysing, reducing, digesting, and desalting the collected plasma samples, the digested peptide segments are labeled using tandem mass tagging (TMT) methods. Afterwards, they are subjected to further fractionation.(23) The re-solubilized peptides were analyzed by liquid chromatography-mass spectrometry/mass spectrometry (LC-MS/MS), and mass spectrometry data were collected. The protein sequences obtained were analyzed using Proteome Discoverer, and selected differential proteins were analyzed for functional enrichment. Detailed procedures are described in the Supplemental data.

### 10× Genomics single-cell RNA sequencing and analysis of PBMCs

Blood samples (5 ml) were collected from two individuals. One individual was a healthy female in her 40s, while the other one was CUA male patient 3 in his 40s. The blood samples of the CUA patient were taken before and after a 3-day treatment involving the use of hAMSCs. PBMC isolation, cell capture, and library construction were performed following the manufacturer’s instructions (10× Genomics). Sequencing was performed on a NovaSeq platform (Illumina). Both the single-cell experiment and sequencing were conducted at the YiKe Population Human Research Institute in Nanjing, China. Each sample was quality controlled, filtered, and standardized for data integration.(24) The obtained datasets were then subjected to dimensionality reduction and clustering for identification of the corresponding cell types. Additionally, functional enrichment analyses were performed. Detailed procedures are described in the Supplemental data.

### Enzyme-linked immunosorbent assay (ELISA)

THBS1 plasma levels were measured in 30 uremic patients, ten CUA patients before hAMSC treatment, and CUA patients 1, 3, 4, 5, 6, 8, and 9 at various time points after hAMSC treatment using the Human Thrombospondin-1 Quantikine ELISA Kit (cat # DTSP10; R&D Systems, Minneapolis, MN, USA). TGF-β1 in plasma was detected using the Human TGF-β1 Quantikine ELISA Kit (cat # DB100C; R&D Systems, Minneapolis, MN, USA), following the manufacturer’s instructions. The standards and the samples were analyzed in duplicate.

### Skin histopathological analysis and immunohistochemistry staining

The skin tissue from the healthy control, CUA patient 3 and the validation cohort before hAMSC treatment, and CUA patient 3 and 4 during hAMSC treatment was obtained through a deep incisional wedge biopsy.(21) The sections were stained with hematoxylin-eosin (HE) and Alizarin red (cat # G1452; Solarbio, Beijing, China) and analyzed using light microscopy. Six-micrometer sections of paraffin-embedded skin tissue were prepared for immunohistochemistry (IHC) analyses. The following primary antibodies were used: mouse anti-THBS1 (cat # ab1823; Abcam, Cambridge, UK) and rabbit anti-CD47 (cat # ab218810; Abcam, Cambridge, UK).

### Statistical analysis

For the analysis of baseline characteristics, measurement data were expressed as mean ± standard deviation or median (interquartile spacing), and categorical data were expressed as proportions or percentages. Independent samples t tests were used to compare normally distributed data between two groups, rank-sum tests were used to compare non-normally distributed data between two groups, and the chi-square test was used to analyze count data. Differences were considered statistically significant when p < 0.05. Statistical analyses were performed using SPSS 22.0 software.

## RESULTS

### Study design and baseline characteristics of the discovery and validation cohorts

Study design is illustrated in Figs. 1A-1C. In the discovery cohort, we performed static plasma in-depth proteomic analysis of uremic and CUA patients and analyzed the dynamic changes in a CUA patient during 15 months of hAMSC treatment at five time points (Fig. 1A). Single-cell transcriptome sequencing of peripheral blood mononuclear cells (PBMCs) was further conducted in a healthy control and a CUA patient treated with hAMSCs (Fig. 1B). In both the discovery cohort (Fig. 1B) and validation cohort (Fig. 1C), the levels of core differentially expressed proteins (DEPs) in the plasma of uremic patients and CUA patients were verified during hAMSC treatment by ELISA. The histopathological features and target proteins were further investigated in the skin tissue of the CUA patients treated with hAMSCs (Fig. 1C). In the discovery cohort (Fig. 1D), the CUA group (n = 3) had significantly higher levels of white blood cells, platelets, albumin, aspartate transaminase and hypersensitive C-reactive protein (hs-CRP) than the uremic group (n = 10). In the validation cohort (Fig. 1E), the CUA group (n=7) had a longer hemodialysis duration and higher levels of white blood cells and hs-CRP compared to the uremia group (n=20).

**Fig 1.**
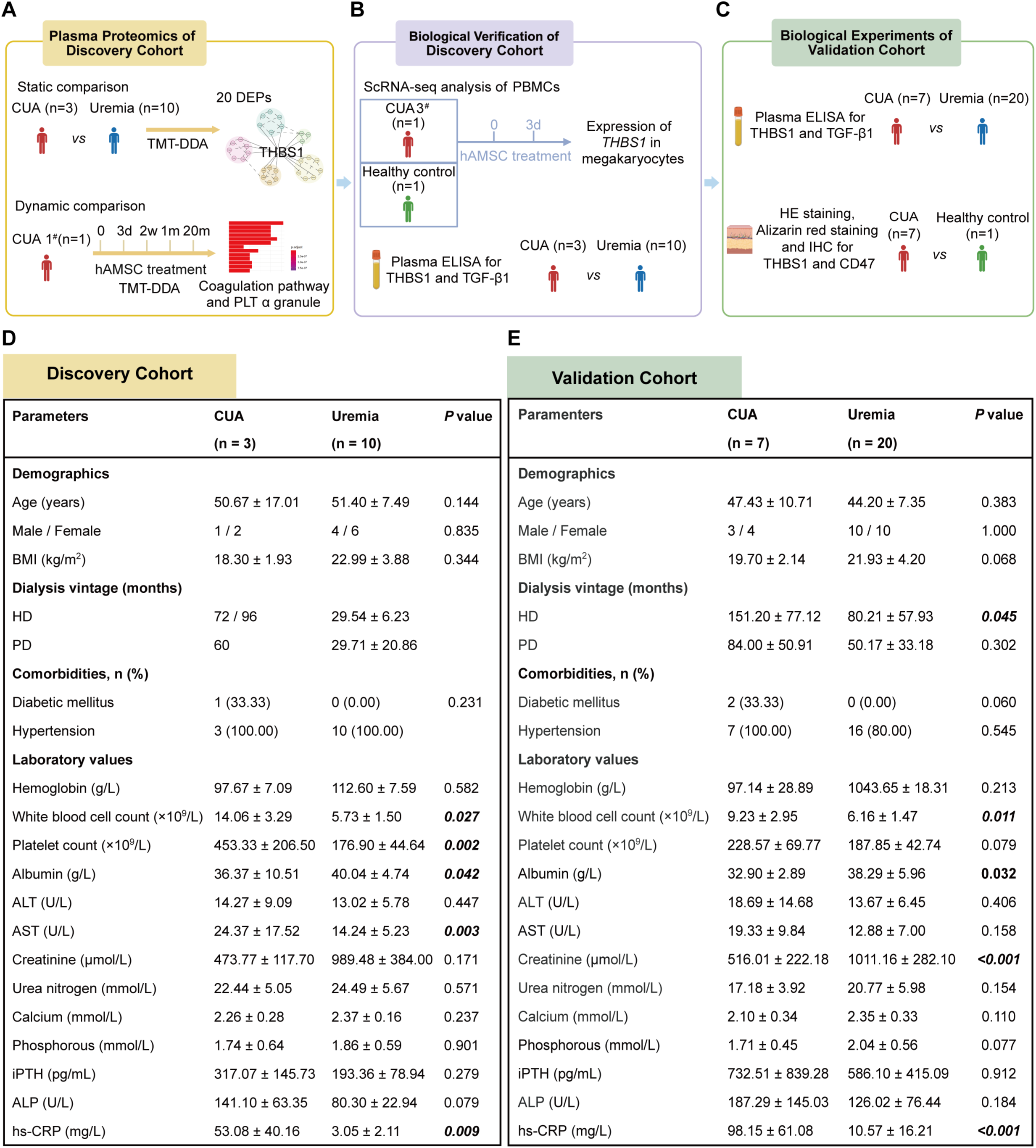
Study design and baseline characteristics of the discovery and validation cohorts. (A-C) Study design. (A) The plasma proteomics of the discovery cohort involved a static analysis of three patients with CUA and ten patients with uremia. Additionally, a dynamic analysis of CUA patient 1 with five time points, who underwent treatment with hAMSCs, was conducted. (B) Biological verification of the discovery cohort: The primary source of increased core DEPs in CUA plasma, THBS1 and TGF-β1, was investigated through single-cell transcriptome sequencing of PBMCs. Additionally, the plasma levels of THBS1 and TGF-β1 in uremia patients and CUA patients treated with hAMSCs were measured using ELISA. (C)The independent validation cohort consisted of 20 patients with uremia and 7 patients with CUA. The plasma levels of THBS1 and TGF-β1 were further investigated by ELISA. Skin histology staining and core DEPs-related immunostaining were also performed. (D-E) Baseline characteristics of demographic and laboratory indicators of the research cohort. (D) The discovery cohort. (E) The validation cohort. CUA calcific uremic arteriolopathy, TMT tandem mass tag-data-dependent acquisition, DEP differentially expressed protein, THBS1 thrombospondin 1, hAMSC human amnion-derived mesenchymal stem cells, ELISA enzyme-linked immunosorbent assay, PLT platelet, PBMCs peripheral blood mononuclear cells, TGF-β1 transforming growth factor-β1, CD47 cluster of differentiation 47, HE hematoxylin-eosin, IHC immunohistochemistry, BMI body mass index, HD hemodialysis, PD peritoneal dialysis, ALT alanine aminotransferase, AST aspartate aminotransferase, iPTH intact parathyroid hormone, ALP alkaline phosphatase, hs-CRP hypersensitive C-reactive protein.

### Rescue of progressive CUA patients with novel hAMSC strategy

CUA patients were followed up from the time they received treatment, with a mean follow-up time of 6.6 months and a maximum time of 20 months. Fig. 2 displays the skin lesions of CUA patients treated with hAMSCs in the discovery cohort. CUA patient 1 was a female in her 30s with rapidly progressing skin lesions within 2 months (Figs. 2A-2C). Skin wound recovery for patient 1 was essentially achieved after the patient underwent hAMSC therapy for 15 months (Figs. 2D-2F). CUA patient 2 was a female in her 60s with skin lesions whose hAMSC treatment was terminated after the first week due to the COVID-19 pandemic (Figs. 2G-2I). CUA patient 3 was a male in his 40s, whose skin wound recovery was achieved after 3 months of hAMSC treatment (Figs. 2J-2L). The HE staining results of the skin tissue from CUA patient 3 indicated that, compared to the tissue necrosis and vascular calcification observed before treatment (Figs. 2M-2N), there was angiogenesis and collagen formation after one month of hAMSC treatment (Fig. 2O).

**Fig. 2.**
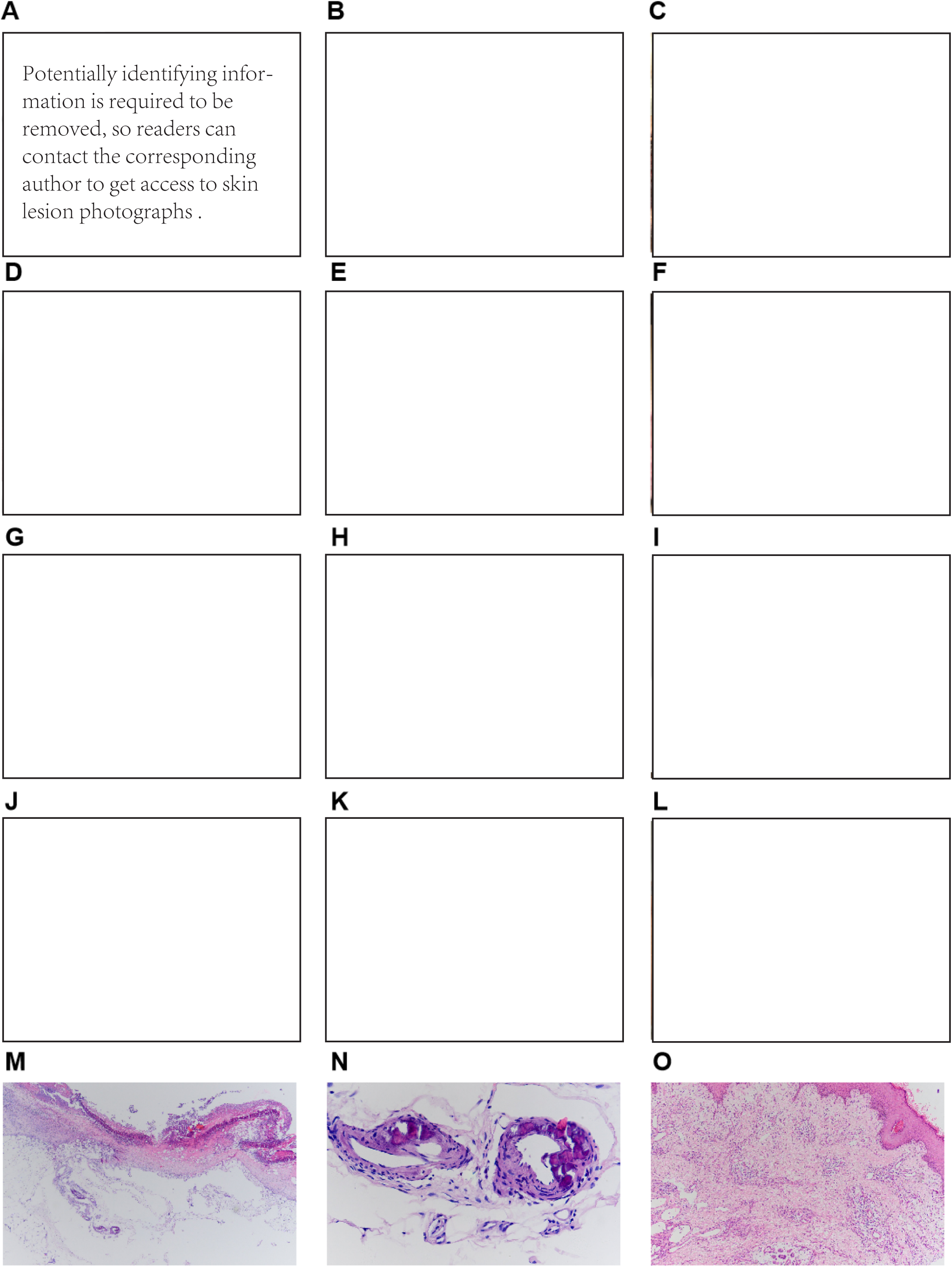
Skin lesions of three calciphylaxis patients treated with hAMSCs in the discovery cohort. (A-C) The rapid progression of thigh skin lesions in calciphylaxis patient 1 before hAMSCs treatment. (A) Erythema, papules and ecchymosis were presented on thigh of both lower limbs with reticulate pattern (July, 2018). (B) Patches of skin necrosis and dark black ecchymosis were seen in large areas of both lower limbs (August, 2018). (C) Progression of lesions with large areas of necrosis on the calf of both lower limbs. Loss of epidermis, covered with black scab, malodorous necrotic ulcer formation in most areas and some reaching the depth of subcutaneous fat (September, 2018). (D-F) After hAMSC treatment, the healing process of skin lesions in calciphylaxis patient 1. (D) Significant improvement of gross ulceration on the calf and the black eschar has been surgically debrided (after hAMSC treatment for 2.5 months). (E) The ulcer area was reduced without exudation, along with small scab formation (after hAMSC treatment for 6 months). (F) The ulcer was nearly healed, only leaving scattered scabs (after hAMSC treatment for 15 months). (G-I) The rapid progression of thigh skin lesions in calciphylaxis patient 2. (G) Irregular erythema and ecchymosis were presented on the thigh (1 week before hAMSC treatment). (H) Progressive necrosis and black eschar on the thigh (on the day of hAMSC treatment). (I) The expanded scope of necrosis and scab on ecchymosis of the thigh (after hAMSC treatment for 1 week). (J-L) The healing process of skin lesions in calciphylaxis patient 3 with hAMSC treatment. (J) The lower limbs showed irregular ulcers with scabs on the edges with exudation (before hAMSC treatment). (K) The area of ulcer and leakage was significantly reduced (after hAMSC treatment for 1 month). (L) The ulcer was totally healed with scar on the leg (after hAMSC treatment for 3 months). (M-O) HE staining of skin biopsy from calciphylaxis patient 3 during hAMSC treatment. (M) The epidermis with large areas of necrosis and crust, reaching the middle and lower layers of the dermis (×40, before hAMSC treatment). (N) Calcium was deposited on the wall of arteriole in subcutaneous fat layer (×400, before hAMSC treatment). (O) The epidermis was healed with hyperplasia of collagen and regenerated microvessels, accompanied by perivascular lymphocyte infiltration (×100, after hAMSC treatment for 1 month).

In the validation cohort, CUA patient 4 was a female in her 30s who underwent a 9-month hAMSC therapy and exhibited complete preservation and regeneration of skin tissue on her lower extremity. CUA patient 5 was a female in her 40s who experienced a noticeable improvement in her clinical manifestations after hAMSC therapy but later discontinued this treatment. CUA patient 6 was a male in his 40s with CKD (stage 3) and a history of parathyroidectomy and kidney transplantation. He received 4 months of hAMSC therapy, which led to clinical improvement in his skin lesions at distal extremities. CUA patient 8 was a female in her 60s with a 20-year history of diabetes. There was a large necrotic black ulcer with purulent discharge and malodor on the right lower limb and foot, and an ulcer was observed on the top of the foot on the left side.

She had received 1 month of hAMSC treatment, but her signs and symptoms improved only mildly due to the large size of her lesions and her poor motivation for further treatment. CUA patient 9 is a male in his 40s who has been on regular hemodialysis for 15 years. He had large, round, deep ulcers on both heels that reached the muscular layer. He had received 2 months of hAMSC treatment and had a significant improvement in bilateral heel lesions and symptomatic relief. CUA patient 10 is a male in his 40s with 19 years of hemodialysis. He developed skin necrosis in both lower limbs, accompanied by the exudation of inflammatory fluid six months before admission to the hospital. They underwent salvage therapy using hAMSCs (except for CUA patients 7 and 10). hAMSC treatment procedures for the patients with CUA (n=8) are provided in detail in Supplemental Table S1.

### The core role of THBS1 in the static plasma proteomic network of CUA *vs* uremic patients

A total of 1619 proteins were identified using a tandem mass tag-data-dependent acquisition mass spectrometry (TMT-DDA MS) workflow (Fig. 3A and Supplemental Table S2). We performed a static proteomic analysis on 20 DEPs (Figs. 3B-3D), followed by an ingenuine pathway analysis (IPA) to explore related pathways (Supplemental Table S3). Compared with the uremic patients, thrombospondin 1 (THBS1 or Tsp-1) was upregulated in the plasma of the CUA group, and the fold change (FC) value of THBS1 was the highest among the DEPs (Fig. 3C). It is noteworthy that THBS1 was the core protein in the static plasma DEP network (Fig. 3E). Furthermore, it was suggested that THBS1 has the effect of inhibiting angiogenesis, as predicted by IPA (Supplemental Table S3).

**Fig. 3.**
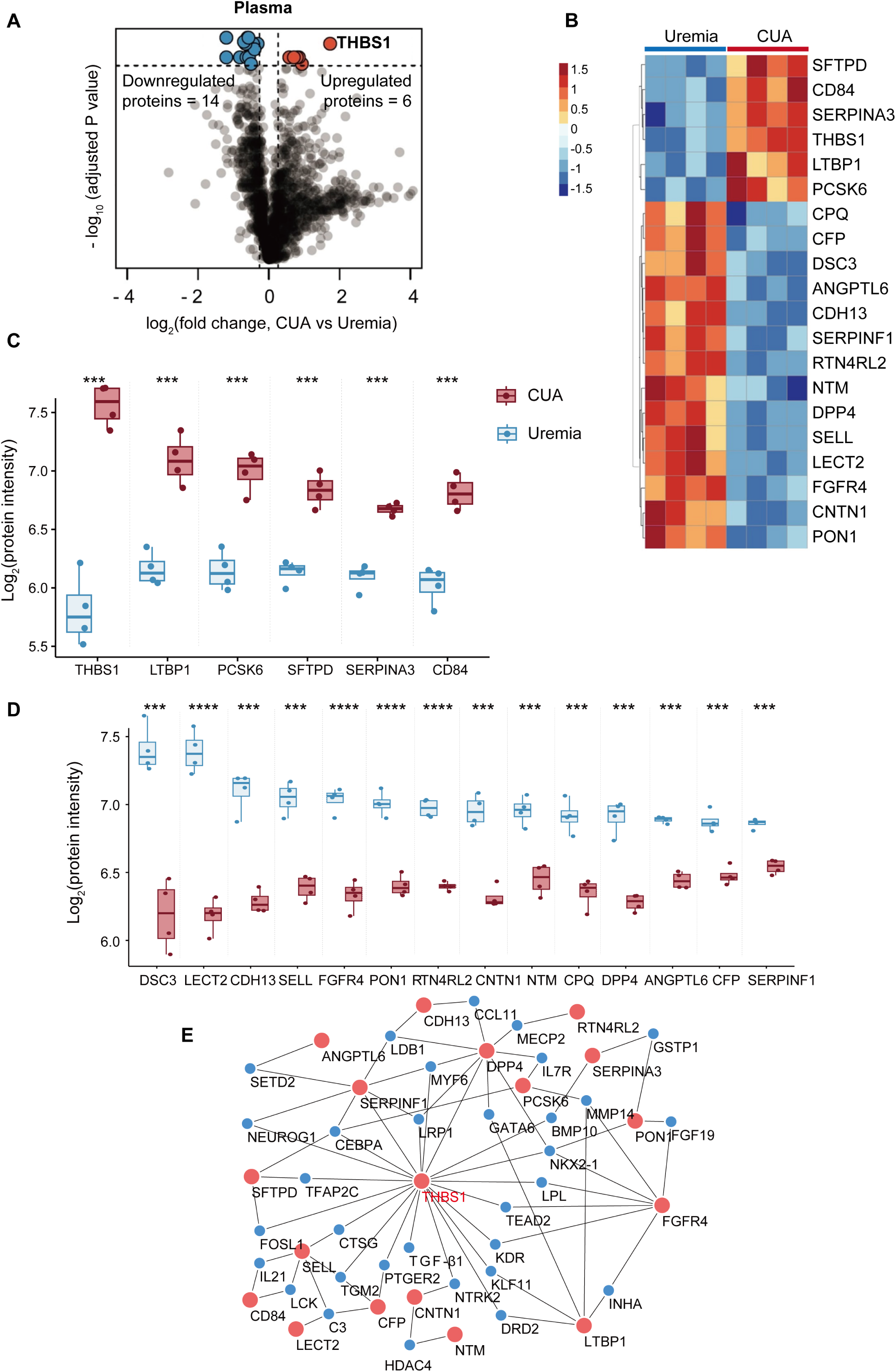
The core role of THBS1 in the plasma DEP network of CUA vs uremic patients in the discovery cohort. (A) The volcano plot of all identified proteins in plasma from CUA patients (n = 3) vs uremic patients (n= 10). Each point represents an individual protein. The horizontal coordinates represent proteins with log2(FC) > 1.2 (red) and log2(FC) < −1.2 (blue). The vertical axis displays −log10(P) > 1.30 (unpaired two-sided Welch’s t test P < 0.05) of DEPs in two technical replicates. In total, 20 DEPs were obtained, including six upregulated and 14 downregulated proteins. THBS1 was upregulated in CUA, and its FC value was the highest among the DEPs. (B) Heatmap showing hierarchical clustering of significant DEPs. (C) Expression of upregulated proteins (n=6) in the discovery cohort. (D) Expression of downregulated proteins (n=14) in the discovery cohort. (F) The interaction network of DEPs was analyzed, with red nodes representing 20 identified DEPs, while blue nodes represent connected proteins predicted by IPA. THBS1 is the core protein of the network. THBS1 thrombospondin 1, DEP differentially expressed protein, CUA calcific uremic arteriolopathy, FC fold change, IPA ingenuine pathway analysis. *** P < 0.001; **** P < 0.0001.

### Process of skin regeneration, dynamic plasma core DEP levels, and enriched pathways in a CUA patient during hAMSC treatment

CUA patient 1 displayed skin regeneration when treated with hAMSCs for 20 months (Fig. 4A). We followed up on CUA patient 1 and tracked changes in the levels of 20 DEPs after treatment with hAMSCs for 3 days, 2 weeks, 1 month, and 15 months (Supplemental Table S4 and Fig. S1). Plasma THBS1 levels were significantly decreased after hAMSC treatment for 3 days. Plasma latent transforming growth factor-beta (TGF-β)-binding protein 1 (LTBP1) exhibited the same trend as THBS1 (Figs. 4B and 4C). We clustered all plasma proteins into eight distinct clusters using Mfuzz (Supplemental Table S5). It was found that proteins in cluster 4 shared the same expression changes over time as THBS1 and LTBP1 (Fig. 4D). We then conducted Gene Ontology (GO) analysis to investigate cluster 4 and found that in the biological process category, these proteins were highly associated with “wound healing”, “hemostasis”, and “blood coagulation”. In the cellular component category, these proteins were associated with “platelet alpha granule” (Fig. 4E and Supplemental Table S6). Together, these results were consistent with the clinical manifestations of skin regeneration (Figs. 2D-2I and Fig. 4A). To account for the potential biological complexity of genes belonging to annotation categories, we developed a protein network (Fig. 4F). We found that THBS1 and TGF-β1 emerged as hub proteins in GO terms related to “wound healing”, “hemostasis”, “blood coagulation”, and “platelet alpha granule”.

**Fig. 4.**
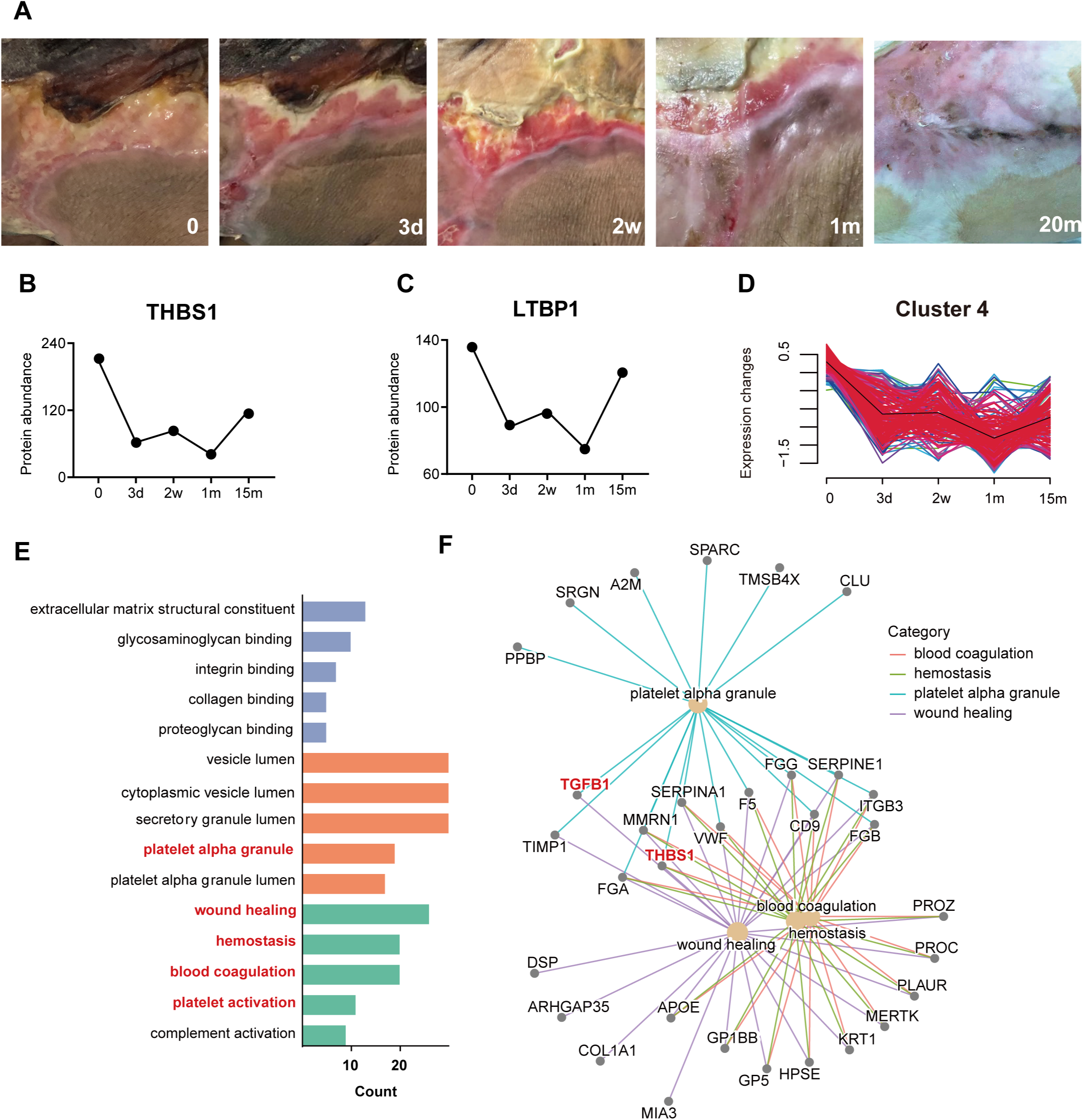
Dynamic characteristics of skin regeneration, core DEPs, and enriched pathways in CUA patient 1 treated with hAMSCs. (A) A significant improvement in the skin lesions of CUA patient 1 was observed following hAMSC therapy. Before treatment, her left hip’s skin showed a typical necrotizing ulcer covered with purulent secretion and black eschar. After 3 days of hAMSC treatment, new granulation tissue was observed extending from the edge of the wound to the center of the ulcer. After 1 month of hAMSC treatment, the epidermis gradually started to regenerate, and the size of the lesions noticeably reduced. After 20 months of treatment, the skin underwent restructuring, forming scar tissue, and eventually fully healed. A fluctuating reduction in plasma THBS1(B) and LTBP1(C) levels in CUA patient 1 was observed after hAMSC treatment. (D) Cluster 4, in which proteins exhibit the same trend as THBS1 and LTBP1 according to Mfuzz. (E) GO analysis of proteins in cluster 4. Significant terms (“wound healing”, “hemostasis”, “blood coagulation”, “platelet alpha granule”) and their enriched proteins, including THBS1 and TGF-β1 were observed in cluster 4 (F). DEP differentially expressed protein, CUA calcific uremic arteriolopathy, hAMSCs human amnion-derived mesenchymal stem cells, THBS1 thrombospondin 1, LTBP1 latent TGF-β binding protein 1, GO Gene Ontology, MF molecular function, CC cellular component, BP biological process.

### Single-cell transcriptome atlas for PBMCs in healthy control and a CUA patient treated with hAMSCs

PBMC samples were derived from CUA patient 3 (before and 3 days after hAMSC therapy) and one healthy control. A total of 12,687 cells passed quality control, and seven cell subtypes were identified, including T cells, B cells, natural killer (NK) cells, monocytes, erythroid-like cells, neutrophils, and megakaryocytes. We visualized the distribution of cells following quality control using Uniform Manifold Approximation and Projection (UMAP), separately by sample group (Fig. 5A). The expression of canonical PBMC genes is shown in Supplemental Fig. S2.

**Fig. 5.**
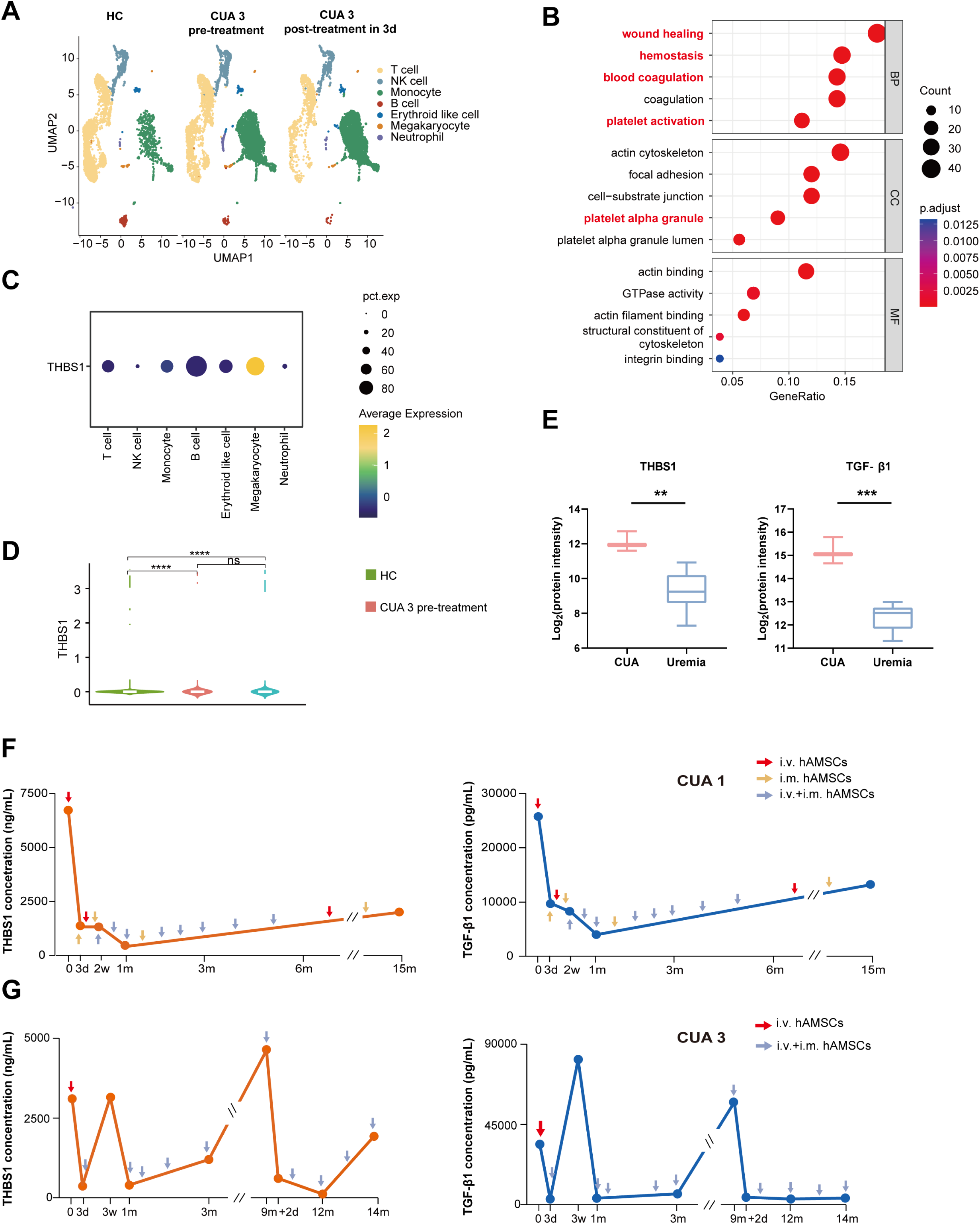
The source, molecular biological function and expression level of THBS1 in CUA revealed by transcriptomics of megakaryocytes and plasma THBS1/TGF-β1 levels during hAMSC treatment. (A) The respective UMAP plots representing the seven clusters across 12,687 PBMCs from two individuals. One individual is a healthy control, while the other is a CUA patient before and after hAMSC treatment for 3 days. (B) Enriched GO terms, including “wound healing”, “hemostasis”, “blood coagulation”, and “platelet alpha granule” for specific genes in megakaryocytes. (C) THBS1 was mainly produced by megakaryocytes, not immune cells in PBMCs. (D) THBS1 levels in each sample. (E) Plasma ELISA measurements of THBS1 and TGF-β1 levels in the discovery cohort. Dynamic plasma levels of THBS1 and TGF-β1 were measured using ELISA in CUA patient 1 (F) and patient 3 (G) during the course of hAMSC treatment. HC healthy control, THBS1 thrombospondin 1, TGF-β1 transforming growth factor-β1, CUA calcific uremic arteriolopathy, hAMSC human amnion-derived mesenchymal stem cell, UMAP uniform manifold approximation and projection, PBMCs peripheral blood mononuclear cells, GO gene ontology, ELISA enzyme-linked immunosorbent assay. ** P < 0.01; *** P < 0.001; **** P < 0.0001.

### *THBS1* was mainly produced by megakaryocytes in the CUA patient

We sought to elucidate the related pathways of megakaryocytes in CUA patient. GO analysis of the specific genes in the megakaryocyte cluster revealed enriched terms, such as “wound healing”, “hemostasis”, “blood coagulation”, and “platelet alpha granule” (Fig. 5B), in agreement with the dynamic plasma proteomics analysis of a CUA patient treated with hAMSCs (Fig. 4E). As shown in Fig. 4B, THBS1 aligned with the cluster trend of Mfuzz 4. This protein plays a significant role in the pathological processes observed in the CUA patient, as well as in regenerative effects resulting from hAMSC treatment (Figs. 5B). We have indicated that megakaryocytes, but not immune cells are the primary source of elevated levels of THBS1 (Fig. 5C). Notably, the expression of THBS1 was significantly higher in CUA patient 3 than in the healthy control (P < 0.01) (Fig. 5D).

### Plasma THBS1 and TGF-β1 levels measured by ELISA in the discovery cohort

Before hAMSC treatment, plasma levels of THBS1 in uremic and CUA patients were 390.02 ± 314.72 ng/ml and 4581.04 ± 1901.60 ng/ml, respectively. Plasma TGF-β1 levels in uremic and CUA patients were 3600.15 ± 1357.17 pg/ml and 38702.26 ± 15924.41 pg/ml, respectively. Compared with the uremic group, the plasma levels of THBS1 and TGF-β1 were increased 11.75- and 10.75-fold, respectively, in the CUA group (Fig. 5E).

The initial intensive hAMSC treatment led to a prompt and substantial decrease in plasma THBS1 and TGF-β1 levels. Nonetheless, in CUA patient 1 over the 15-month follow-up period, while these levels continued to be relatively low, there was a gradual rise as the frequency of hAMSC therapy decreased (Fig. 5F). In CUA Patient 3, the plasma showed low levels of THBS1 and TGF-β1 after hAMSC treatment. Consequently, the patient discontinued treatment for a period of six months. However, upon resuming hAMSC treatment, both THBS1 and TGF-β1 were initially found to be present at high levels, only to decrease with further hAMSC therapy (Fig. 5G).

### THBS1-related biological experiments were conducted on plasma and skin tissue samples from the verification cohort

HE and Alizarin red staining for skin tissues from the CUA validation cohort is shown in Figure 6A-6C and 6E-6L, with marked inflammatory cell infiltration, necrosis, microvascular calcification, and microthrombosis. Fig. 6D indicates significant improvement in skin histopathology after 1 month of hAMSC treatment in CUA patient 4. Compared with the pre-treatment CUA group, the plasma levels of THBS1 and TGF-β1 showed a remarkable decrease, respectively, in the uremia group and post-treatment CUA group (Fig. 6M). The dynamic plasma levels of THBS1 and TGF-β1, measured using ELISA, were depicted in Supplemental Fig. S3 for CUA patients 4, 5, 6, 8, and 9 during the period of hAMSC treatment. After 3 days of hAMSC therapy, the levels of plasma THBS1 and TGF-β1 in the five CUA patients decreased by 9.75- and 4.59-fold, respectively (Supplemental Fig. S4). Due to the interaction between inflammation and THBS1, we also compared the blood hs-CRP levels of CUA patients treated with hAMSCs before and 3 days after treatment and found no statistically significant difference (Supplemental Fig. S5).

**Fig. 6.**
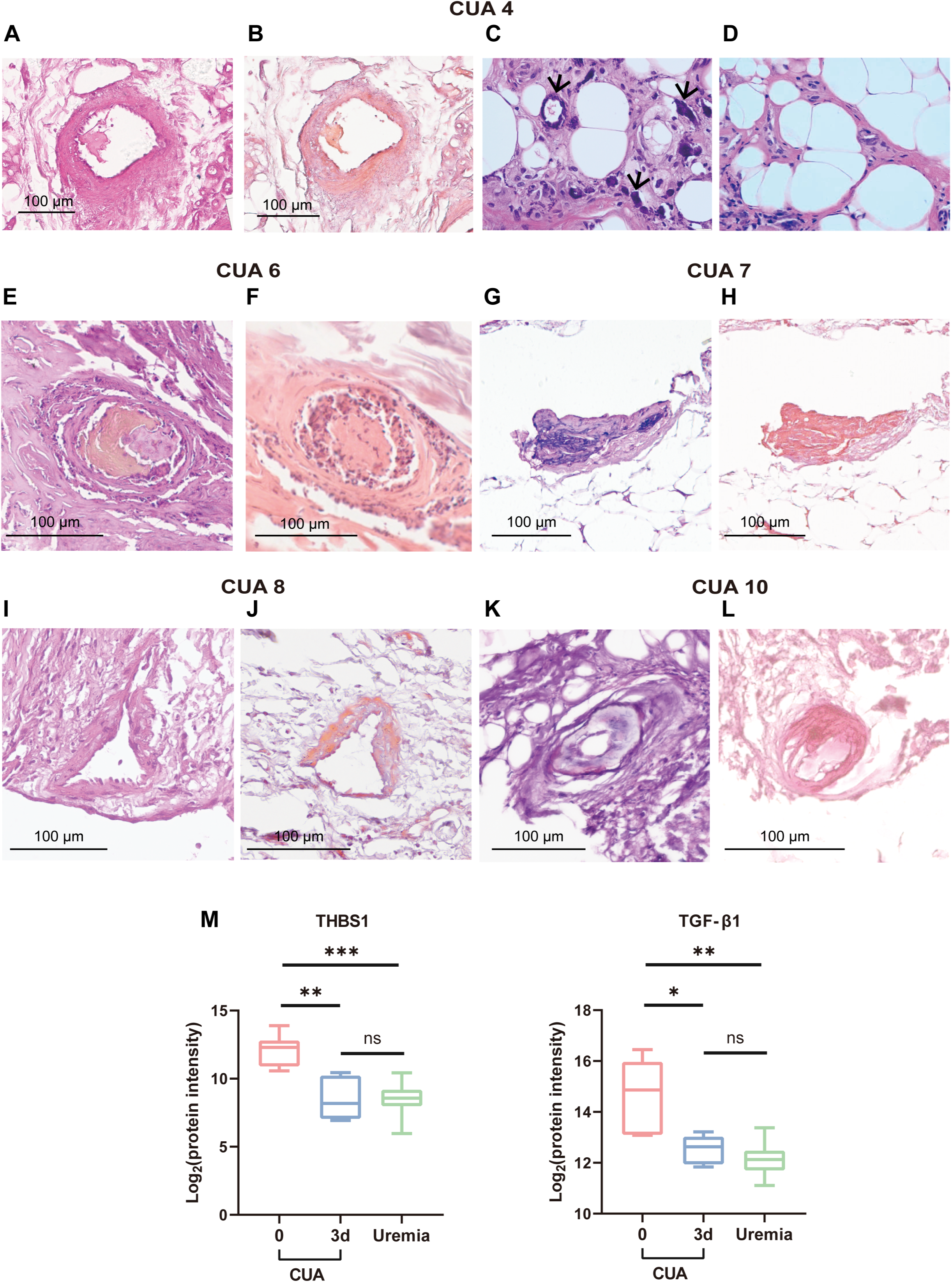
Diagnosis and biological experiments of the independent CUA validation cohort. (A-L) HE and Alizarin red staining of skin tissue in five patients. (A-B) CUA patient 4. HE staining indicated necrosis and thrombosis in the microvascular (A). ARS displayed annular calcification of the vessels and calcium deposition (B). (C-D) Skin samples from CUA patient 4 before and 1 month after hAMSC treatment (HE staining, × 400). Calcium deposition in microvessels and adipose areas 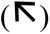 before hAMSC treatment (C). Decreased calcium deposition around microvessels and adipose tissue after 1-month hAMSC therapy (D). (E-F) Skin samples from CUA patient 6 revealed inflammation, tissue necrosis and microthrombosis (E, HE staining) and microvascular calcification (F, ARS). (G-H) Skin samples from CUA patient 7 indicated calcification and necrosis in microvasculature by HE (G) and ARS (H). (I-J) Skin samples from CUA patient 8 revealed inflammation, tissue necrosis, microvascular calcification, and calcium deposition by HE (I) and ARS (J). (K-L) Skin samples from CUA patient 10. HE staining revealed inflammation, swelling and necrosis of the microvessel (K), with microvascular calcification (ARS, L). (M) The plasma levels of THBS1 and TGF-β1 in the uremic patients (n=20), the CUA patients before (n=7), and after hAMSC treatment for 3 days (n=5). HE hematoxylin-eosin, ARS Alizarin red staining. * P < 0.05; ** P < 0.01; *** P < 0.001.

Skin samples from the healthy control showed collagen fibers, intact blood vessels and no expression of THBS1 and CD47 (Figs. 7A-7C). Skin samples from CUA patient 5 before hAMSC treatment revealed a microthrombus filling the arteriole (Fig. 7D). THBS1 and its receptor, CD47, were predominantly expressed at the site of thrombosis (Figs. 7E-7F). Skin samples from CUA patient 9 before hAMSC treatment suggested thrombogenesis in the capillaries and infiltration of inflammatory cells (Fig. 7G). Alizarin red staining displayed annular calcification of the vessels and calcium deposition within the thrombus (Fig. 7H). THBS1 and CD47 were mainly expressed in these thrombi (Figs. 7I-7J).

**Fig. 7.**
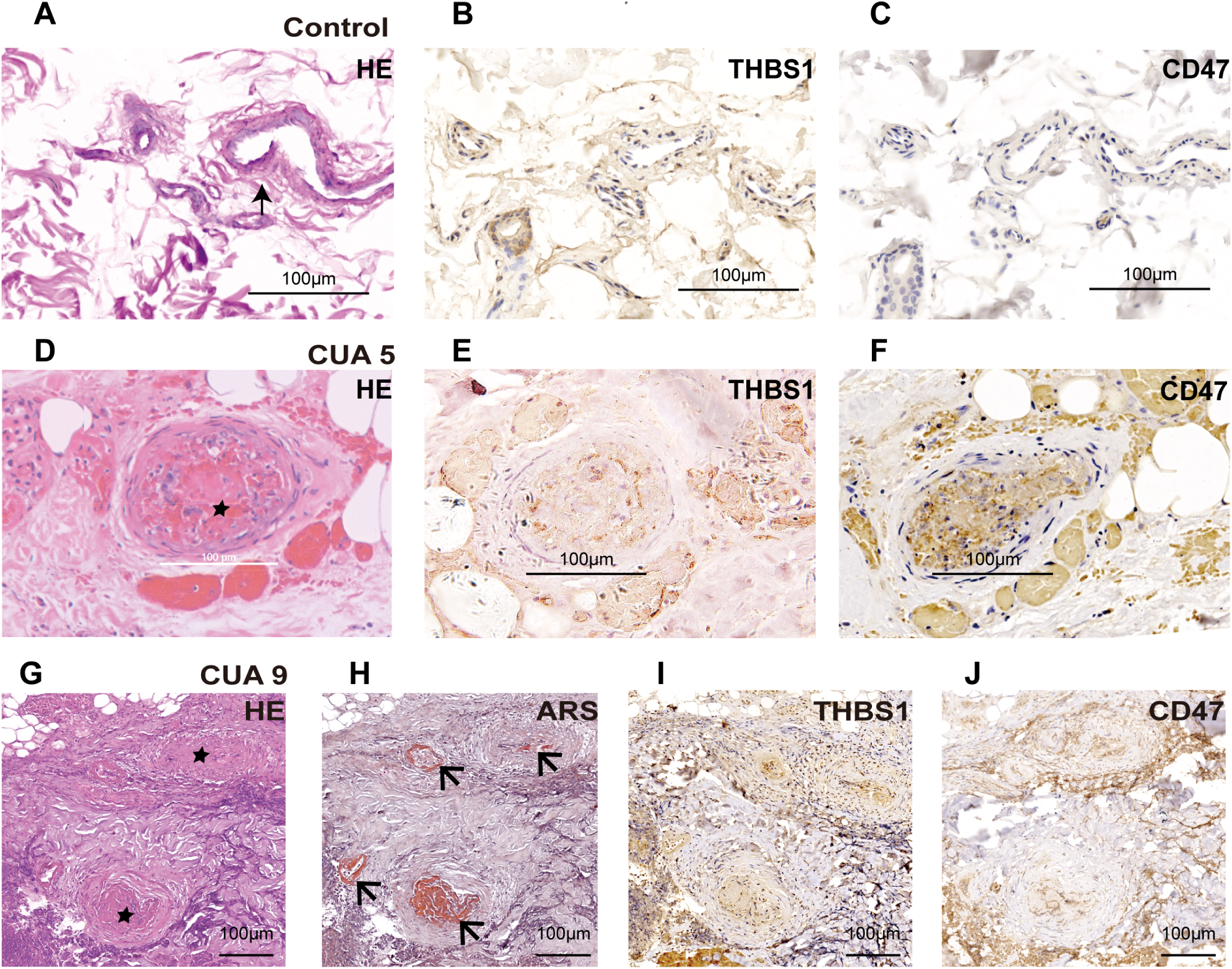
Histopathological analysis of biomarker-related proteins in skin tissue from the healthy control and independent CUA validation cohort. (A-J) Histopathological analysis was conducted to identify target proteins in the skin. Hematoxylin-eosin staining and immunohistochemical analyses of THBS1 and CD47 were conducted in the skin tissue from the healthy control and CUA patient 5 before hAMSC treatment (A-F). Pathological characteristics of the healthy control included intact blood vessels 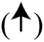, and no expression of THBS1 and CD47 (A-C). Skin samples from CUA patient 5 before hAMSC treatment revealed a microthrombus 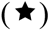 filling the arteriole, at which site THBS1 and CD47 were expressed (D-F). Skin tissue from CUA patient 9 was analyzed by HE staining, Alizarin red staining, and immunohistochemistry for THBS1 and CD47 (G-J). HE staining suggested thrombogenesis 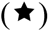 in the capillaries (G). Alizarin red staining displayed annular calcification 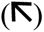 of the vessels and calcium deposition 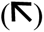 within the thrombus (H). THBS1 and CD47 were found in these thrombi (I-J). THBS1 thrombospondin 1, TGF-β1 transforming growth factor-β1, CUA calcific uremic arteriolopathy, hAMSC human amnion-derived mesenchymal stem cell, CD47 cluster of differentiation 47, HE hematoxylin-eosin, ARS Alizarin red staining.

## DISCUSSION

Our research indicates that hAMSC therapy holds promise for calciphylaxis patients. However, the key challenge lies in developing non-invasive diagnostic methods and establishing individualized precision treatment strategies. In the present study, we conducted a static and dynamic plasma proteomic analysis for CUA patients treated with hAMSCs.(23, 25) Compared to the uremic group, THBS1 was the core upregulated DEP in CUA patients. The plasma levels of THBS1 promptly and significantly decrease after hAMSC treatment. Plasma LTBP1 levels show the same fluctuating trend as THBS1.

THBS1 is released by activated platelet alpha granules, with physiological levels in healthy adult plasma typically ranging from 20 to 40 ng/ml.(26) THBS1 can be found abundantly in both megakaryocytes and platelets. Under conditions of injury and stress, THBS1 can also be produced by innate immune cells,(27) endothelial cells, smooth muscle cells, etc..(28) As a 420-kDa multidomain glycoprotein, THBS1 cannot be cleared by dialysis (29) and has multiple functions, including enhancing platelet aggregation,(30) thrombin stimulation,(31) inducing endothelial cell apoptosis,(32) mediating vascular injury, affecting angiogenesis,(33) enhancing systemic inflammation (34) and regulating the immune response.(35)

Although named calciphylaxis, its intricate clinical manifestations cannot be completely explained by vascular calcification, which is a common complication in patients with CKD. The rapid progression of calciphylaxis and its high mortality rate have been emphasized. These can be elucidated through the biological functions of THBS1. The CUA patients did not adjust the use of anticoagulant drugs during the hAMSC treatment period within one week. Additionally, the decrease in blood hs-CRP levels in CUA patients three days after treatment was less pronounced than that of THBS1 when compared to pre-treatment levels. These results indicate that the use of anticoagulant drugs and inflammatory-related indicator (hs-CRP) are not confounding factors for THBS1 as a potential blood biomarker for CUA patients.

TGF-β1 usually exists as an inactive large latent complex formed by the covalent binding of the C-terminal mature TGF-β1 peptide and the N-terminal latency-associated peptide to LTBP1 through disulfide bonds. (36) THBS1 binds to the latent TGF-β complex through the Lys-Arg-Phe-Lys sequence in the type 1 repeat,(37) and induces activation by stimulating conformational changes in the latent complex.(38) This process is involved in biological processes such as wound healing, cell proliferation,(39) the inflammatory response, and fibrosis.(40)

In our plasma proteomic data, the GO analysis represented terms including “wound healing,” and “platelet alpha granule,” with enriched proteins THBS1 and TGF-β1. Based on our PBMC transcriptomic data, THBS1 primarily originated from megakaryocytes, the primary producers of platelets,(41) rather than from immune cells, in CUA patient 3. These findings support the notion that the increased production of THBS1 primarily originates from platelets in CUA patients.

ELISA measurements further confirmed that plasma levels of THBS1 and TGF-β1 were significantly higher in the CUA group compared to the uremic group. Despite displaying a rapid and significant reduction, a gradual increase in THBS1 and TGF-β1 levels was observed during the follow-up period as the frequency of hAMSC therapy decreased. It is crucial to closely monitor the levels of THBS1 and TGF-β1 throughout the treatment period, particularly in the initial stage. Prompt adjustments to the frequency of hAMSC therapy must be made to maintain low plasma levels of THBS1 and TGF-β1.

The pathogenesis of calciphylaxis remains poorly understood. However, there is emerging evidence suggesting that hypercoagulability may contribute to its development.(42) Vascular calcification is considered the primary pathological process in the initiation of calciphylaxis lesions. The underlying hypercoagulable state may contribute to intravascular thrombosis in pre-existing arteriole injuries.(43) As a result, this can lead to cutaneous necrosis and ischemic pain. By binding to the cell surface receptor CD47, THBS1 can block endothelial cell proliferation,(44) produce oxidative stress,(45, 46) induce apoptosis, and limit blood flow.(28) THBS1-CD47 signaling also contributed to platelet activation and thrombus formation.(47) Our immunostaining data from the skin tissue of CUA patients revealed increased expression of THBS1 and CD47, especially at the site of microthrombosis.

This study has several limitations. First, the heterogeneity of CUA skin lesions and infection status, along with the COVID-19 pandemic, has resulted in inconsistent dosages of hAMSC treatment and follow-up care for CUA patients. Second, due to the rarity of calciphylaxis,(1) we have to face the challenge of limited sample size. Although the sample size of rare diseases is usually small,(48, 49) it will be imperative to recruit a larger number of calciphylaxis patients at multiple centers, optimize treatment protocols and follow-up procedures, thereby enhancing the broad applicability and persuasiveness of our findings.

In conclusion, our multidimensional exploration with multiple tissues provides a novel perspective on the acute impacts of progressive vascular injury, which has been highlighted as a key factor associated with skin tissue necrosis in patients with CUA. Elevated plasma THBS1 and TGF-β1 levels can be rapidly and dramatically reduced by intensive hAMSC therapy. THBS1 and TGF-β1 deserve further investigation as blood-based potential prognostic biomarkers for this disastrous disease, optimizing hAMSC therapeutic strategy.

## DATA AVAILABILITY

The proteomic data has been deposited in iProX (IPX0009031001, https://www.iprox.cn/page/DSV021.html;?url=1718120623768Uh8P, password h84O) and will be made publicly available upon publication of this manuscript. The single-cell RNA sequencing data has been deposited in NGDC (OMIX006677, https://ngdc.cncb.ac.cn/omix/preview/iuik4gl4. No customized code in proteomics analysis. The single-cell transcriptome analysis code used in this study has been deposited at GitHub under https://github.com/lsj46/CUA-Code/blob/master/Code.Rmd. However, raw data for individual patients cannot be publicly disclosed due to patient confidentiality reasons. Researchers can apply for access to these data, subject to approval from the individual institutional review boards. Any additional information required to reanalyze the data reported in this work paper is available from the lead contact upon request.

## Supplemental data

This article contains supplemental data. ((24, 50–57))

## Acknowledgments

The authors would like to thank all of the study participants and their families, study coordinators, and support staff involved in the rescue of patients with calciphylaxis for making this study possible. This work was funded by the National Natural Science Foundation of China (81270408, 81570666, 81730041, and 81671447), National Key R&D Program of China (2022YF0608403, 2021YFA1301600), the International Society of Nephrology (ISN) Clinical Research Program (18-01-0247), CKD Anemia Research Foundation from China International Medical Foundation (Z-2017-24-2037), Natural Science Foundation of Jiangsu Province (BK20243054), Outstanding Young and Middle-Aged Talents Support Program of The First Affiliated Hospital of Nanjing Medical University (Jiangsu Province Hospital), Jiangsu Province Hospital (the First Affiliated Hospital with Nanjing Medical University) Clinical Capacity Enhancement Project (JSPH-MA-2023-7), the National Key Research and Development Program of China (2017YFC1001303), the State Key Laboratory of Reproductive Medicine Program (SKLRM-GC201803), the State Key Laboratory of Reproductive Medicine and Offspring Health Program (SKLRM-K202105), “Pioneer” and “Leading Goose” R&D Program of Zhejiang (2024SSYS0035) and Westlake Omics Junior Clinician Support Program 2021. The study was supported by the ISN Mentorship Program and the authors thank Professor Marcello Tonelli (University of Calgary, Canada) for his helpful comments on the draft of the manuscript. The authors thank Dr Liang Chen, from the School of Medicine at Westlake University, for his guidance in drawing the images. The authors thank Dr Juncheng Dai from the School of Public Health at Nanjing Medical University for his guidance in analysing the single-cell transcriptome data. The authors thank Drs Renqiu Wang and Min Dong at Department of Nephrology, ChuiYangLiu Hospital Affiliated to Tsinghua University for helping to rescue the CUA patient. The authors thank Dr Guangquan Xun at Department of Orthopaedics, Yingkou Yanghe Hospital for providing blood and skin tissue samples. We thank LetPub (www.letpub.com) for its linguistic assistance during the preparation of this manuscript.

## Author contributions

Project conceived and directed, hAMSC clinical treatment design, and research funding support by Ningning Wang, Jiayin Liu, Tiannan Guo, Yi Zhu, Lianju Qin; research ethical issues and administrative guidance by Xiuqin Wang and Ningxia Liang; hAMSC research platform management, hAMSC preparation, cell line construction, quality control by Jiayin Liu, Lianju Qin, Yugui Cui, Chunyan Jiang, Xiang Ma and Song Ning; the clinical research design was guided by Feng Chen and Shaowen Tang; clinical management and multidisciplinary team rescue of the CUA patients by Ningning Wang, Changying Xing, Ming Zeng, Chun Ouyang, Jingjing Wu, Kang Liu, Yanggang Yuan, Hongqing Cui, Ling Zhang, Yongwu Yu, Haibin Ren, Li Zhang; skin tissue histological analysis and immunostaining by Zhonglan Su, Jingfeng Zhu, Ningning Wang, Youjia Yu, Cui Li, Shijiu Lu, Jiaying Hu, Guicun Fang and Meihua Liao; follow-up study by Ningning Wang, Xiaoxue Ye, Shijiu Lu, Jing Zhang, Cui Li, Daoyu Wu; clinical specimen collection, measurement, and data acquisition by Ningning Wang, Xiaoxue Ye, Shijiu Lu, Jing Zhang, Ming Zeng, Zhanhui Gao, Xinfang Tang, Baiqiao Zhao, Anning Bian, Fan Li, and Cuiping Liu; proteomics and single-cell transcriptomics sample preparation and analysis by Tiannan Guo, Yi Zhu, Yaoting Sun, Xiaoxue Ye, Lu Li, Ningning Wang, Jing Zhang, Weigang Ge, and Xiying Mao; image creation and modification by Xiaoxue Ye, Ningning Wang, Shijiu Lu, Jiaying Hu; interpretation of the data, writing, submission and revision of the manuscript by Xiaoxue Ye, Ningning Wang, Yi Zhu, Tiannan Guo, Shijiu Lu, Yaoting Sun, Lianju Qin, and Jing Zhang. All other authors reviewed the final draft and agreed to publish the manuscript.

## Conflict of interest

The authors declare no competing interests.

## Abbreviations

CUA, calcific uremic arteriolopathy; hAMSCs, human amnion-derived mesenchymal stem cells; DEPs, differentially expressed proteins; THBS1, thrombospondin 1; TGF, transforming growth factor; LTBP1, Latent transforming growth factor-β binding protein 1; PBMCS, peripheral blood mononuclear cells; ELISA, enzyme-linked immunosorbent assay; CKD, chronic kidney disease; HD, hemodialysis; STS, sodium thiosulfate; SHPT, secondary hyperparathyroidism; PTH, parathyroid hormone; TMT, tandem mass tagging; LC-MS/MS, liquid chromatography-mass spectrometry/mass spectrometry; HE, hematoxylin-eosin; IHC, immunohistochemistry; hs-CRP, hypersensitive C-reactive protein; DDA, data-dependent acquisition; IPA, ingenuine pathway analysis; FC, fold change; GO, Gene Ontology; NK, natural killer; UMAP, Uniform Manifold Approximation and Projection; PLT , platelet; CD47; cluster of differentiation 47; BMI, body mass index; PD, peritoneal dialysis; ALT, alanine aminotransferase; AST, aspartate aminotransferase; ALP, alkaline phosphatase; MF, molecular function; CC, cellular component; BP, biological process; ARS, Alizarin red staining.

## Notes

### Competing Interest Statement

The authors have declared no competing interest.

### Clinical Trial

NCT04592640

### Author Declarations

This study was approved by the ethics committee of the First Affiliated Hospital of Nanjing Medical University in China (2018-QT-001, 2020-QT-01, 2020-QT-09, 2020-12-02).

