## Supplementary material for "Multi-omics Analysis Reveals Prognostic Biomarker Candidates for Calcific Uremic Arteriolopathy Patients Treated with Stem Cells": Description of Additional Supplemental Table.docx

**Title: Supplemental Table S1.**

**Description:** Description of hAMSC treatment procedures for eight CUA patients.

hAMSCs were administered intravenously to the patient at a dosage of 1.0 × 10^6^ cells/kg body weight, combined with local intramuscular injection along the wound edge (2.0 × 10^4^ cells/cm^2^) and external application of the cell culture supernatants on wound surface.

Abbreviations: hAMSC, human amniotic mesenchymal stem cell; CUA, calcific uremic arteriolopathy; IVD, intravenous drip; IM, intramuscular injection.

**Title: Supplemental Table S2.**

**Description:** Plasma proteomic matrix of uremic patients (n=10) and CUA patients (n=3) in the discovery cohort.

Abbreviations: CUA, calcific uremic arteriolopathy.

**Title: Supplemental Table S3.**

**Description:** Twenty differentially expressed plasma proteins and their IPA analysis of CUA patients were compared to uremic patients in the discovery cohort.

Sheet 1, Plasma differentially expressed proteins in uremic patients with or without CUA; Sheet 2, IPA molecule analysis of plasma differentially expressed proteins in uremic patients with or without CUA; Sheet 3, IPA pathways analysis of plasma differentially expressed proteins in uremic patients with or without CUA.

Abbreviations: IPA, Ingenuine pathway analysis; CUA, calcific uremic arteriolopathy.

**Title: Supplemental Table S4.**

**Description:** Plasma proteomic matrix of CUA patient 1 during the course of hAMSC treatment.

There are five time points: before and after hAMSC treatment at 3 days, 2 weeks, 1month, and 15months.

Abbreviations: CUA, calcific uremic arteriolopathy; hAMSC, human amniotic mesenchymal stem cell.

**Title: Supplemental Table S5.**

**Description:** Diverse dynamic changes of plasma protein levels were observed in CUA patient 1 at different time points during hAMSC treatment, as clustered by Mfuzz.

Abbreviations: CUA, calcific uremic arteriolopathy; hAMSC, human amniotic mesenchymal stem cell.

**Title: Supplemental Table S6.**

**Description:** GO analysis of Mfuzz 4 resulted in a series of plasma proteins with the same trendline of THBS1 at different time points, since CUA patient 1 received hAMSC treatment.

Abbreviations: GO, gene ontology; CUA, calcific uremic arteriolopathy; hAMSC, human amniotic mesenchymal stem cell.
