## Supplementary material for "Multi-omics Analysis Reveals Prognostic Biomarker Candidates for Calcific Uremic Arteriolopathy Patients Treated with Stem Cells": Supplementary Information_Clean Version.pdf

Supplementary methods

Supplementary references

Supplementary Fig. 1: Dynamic trends of 20 differentially expressed proteins in the plasma of CUA patient 1 treated with hAMSCs were analyzed through proteomic analysis.

Supplementary Fig. 2: Cell type identification in the scRNA-seq integrated datasets of PBMCs from a healthy control and CUA patient 3.

Supplementary Fig. 3: Dynamic THBS1/TGF-β1 plasma levels of CUA patients in the validation cohort were measured by ELISA during the course of hAMSC treatment.

Supplementary Fig. 4: Plasma THBS1 and TGF-β1 levels of five CUA patients were measured in the validation cohort before and after treatment with hAMSCs for 3 days.

Supplementary Fig. 5: Comparison of hs-CRP levels before and 3 days after hAMSC treatment in CUA patients.

### Supplementary methods

#### Protein digestion

High-abundance proteins in plasma samples were depleted by of High-Select™ Top14 abundant protein depletion resin (Thermo Fisher Scientific). The plasma samples were then transferred to Microcon centrifugal filters with a molecular weight cut-off of 10 kDa for protein concentration. Next, the samples were lysed with a lysis buffer (6 M urea (Sigma-Aldrich, Cat # U1250) and 2 M thiourea (Sigma-Aldrich, Cat # T8656) resolved in triethylammonium bicarbonate buffer (Sigma-Aldrich, Cat # T7408). The proteins were reduced with 10 mM tris (2-carboxyethyl) phosphine (TCEP, Adamas-beta, Cat # 61820E) and alkylated with 40 mM iodoacetamide (IAA, Sigma-Aldrich, Cat # I6125) in darkness [1]. Furthermore, the proteins were digested with lysine protease (LysC, Hualishi Tech, Cat # HLS LYS001C) for 4 h and with trypsin (Hualishi Tech, Cat # HLS TRY001C) for 8 h, with an enzyme-to-substrate ratio of 1:40 for each step. The reaction was stopped by 10% trifluoroacetic acid (TFA, Thermo Fisher Scientific, Cat # 85183). Peptides were desalted using SOLAµ (Thermo Fisher Scientific, Cat # 62209-001).

Cleaned peptides were labeled with TMTpro 16plex label reagents (Thermo Fisher Scientific, Cat # A44520). High-pH fractionation was performed using a 120-min gradient on an XBridge BEH130 C18 Peptide Separation Technology (PST) column (300 Å, 5 µm, 4.6 mm × 250 mm, 1/pk; Waters Corporation, MA, USA) connected to a nanoflow DIONEX Ultimate 3000 system (Thermo Fisher Scientific™, San Jose, USA) [2]. The gradient was from 5% to 35% acetonitrile (ACN, Thermo Fisher Scientific, Cat # A955-4) with pH 10.0 at a flow rate of 1 ml per minute, and 60 fractions were collected equidistantly, which were finally combined into 30 fractions.

#### Mass spectrometry data acquisition

The redissolved peptides were analyzed by liquid chromatography (LC)-MS/MS with the same LC system coupled to an Orbitrap Exploris 480 mass spectrometer (Thermo Fisher Scientific™, San Jose, USA), equipped with a FAIMS Pro™ (Thermo Fisher Scientific™, San Jose, USA) in data-dependent acquisition (DDA) mode.

For each acquisition, peptides were loaded onto a precolumn (3 µm, 100 Å, 20 mm × 75 µm i.d.) using a 60-min LC gradient (from 7% to 30% buffer B) at a flow rate of 300 nl/min (analytical column, 1.9 µm, 120 Å, 150 mm × 75 µm i.d.). Buffer A was 2% ACN with 98% $H_2O$ containing 0.1% formic acid (FA, Thermo Fisher Scientific, Cat # A117-50), and buffer B was 98% ACN with water containing 0.1% FA. All reagents were MS grade. The *m/z* range of MS1 was 375-1800, the resolution at full width at half maximum (FWHM) was 60,000, the normalized automatic gain control (AGC) target was 300% with an intensity threshold of 2e4, and the maximum ion injection time (max IT) was 50 ms. MS/MS experiments were performed with a resolution at FWHM of 30,000, a normalized AGC target of 200%, and a max IT of 86 ms. The turbo-TMT and advanced Peak Determination were enabled, the isolation window was set to 0.7 Da, and the first mass was set to 110 *m/z*.

**Proteomic data processing**

The resultant mass spectrometric data were analyzed using Proteome Discoverer (Version 2.4.1.15, Thermo Fisher Scientific, San Jose, USA) against the FASTA downloaded from Human SwissProt on 15 July 2020, containing 20,368 reviewed protein sequences.

**Identification of proteins and specific clusters**

From all identified proteins in plasma from uremic calciphylaxis patients (n = 3) vs paired non-calciphylaxis patients (n = 10), DEPs were selected by the unpaired two-sided adjusted Welch's t test ($p < 0.05$; fold change > 1.2 or fold change < −1.2). We clustered the plasma proteins of CUA patient 1 after hAMSC treatment using Mfuzz into eight significant discrete clusters respectively. THBS1 was classified into cluster 4.

**Pathway analysis**

Network pathway analysis tools were used for proteins in cluster 4. The GO processes were enriched by R and visualized as a classical pathway map, enrichment pathways, and a differential protein network relationship map. IPA of the regulated proteins identified the most significant pathways with the P value determined based on a right-tailed Fisher's exact test with the overall activation or inhibition states of enriched pathways predicted by z-score [3].

### PBMCs dissociation and preparation

The PBMCs were isolated by density gradient centrifugation using Ficoll-Paque™ PLUS medium (Cytiva, Cat # 17144003) and washed with Ca/Mg-free PBS (Thermo Fisher Scientific, Cat # 14040141). To remove the red blood cells, 2 mL red blood cell lysis buffer (Roche, Cat # 11814389001) was added at 25°C for 10 minutes. The solution was then centrifuged at 500 × g for 5 minutes and suspended in PBS. The blood samples were centrifuged at 400g for 5 minutes at 4°C, and the supernatant was discarded. After removing the red blood cells, PBMCs were isolated by centrifuging at 400g for 10 minutes at 4°C. The supernatant was discarded, and the PBMCs were resuspended to obtain a single-cell suspension.

### Single-cell RNA sequencing

Single-cell suspensions were converted into barcoded scRNA-seq libraries by using the Chromium Next GEM Single Cell 3′ GEM, Library & Gel Bead Kit v3.1 (10× Genomics, Cat # 1000121), following the manufacturer's instructions. Libraries were prepared using 10× Genomics Library Kits and sequenced on the Illumina Nova6000 with 150 bp paired-end reads.

### Single-cell RNA-seq data processing

Quality control was applied to cells based on three specific metrics. Firstly, the total unique molecular identifier (UMI) count was considered. Cells with less than 1000 UMIs were excluded. Secondly, the number of detected genes in each cell was examined. Cells with a low number of detected genes were also excluded. Lastly, the proportion of sequences mapped to mitochondrial genes was evaluated. Cells with more than 20% of sequences mapped to mitochondrial genes were excluded from the analysis. We employed the deconvolution strategy implemented in the R package scran to normalize the UMI counts. Specifically, we computed cell-specific size factors using the computeSumFactors function, which were subsequently used to scale the counts for each cell. The resulting logarithmic normalized counts were then utilized for the downstream analysis.

**Single-cell normalization and dataset integration**

The raw UMI counts for the three samples were normalized and integrated using Seurat (v4.3.1) [4]. Unless stated otherwise, all analyses were performed using default parameters. Cells with fewer than 200 non-zero genes and genes found in fewer than 10 cells were excluded. To mitigate the effects of low-quality cells [5], each sample underwent a manual assessment based on standard quality control parameters. These parameters included the total number of genes per cell, the total number of unique genes, and the fraction of sequences mapping to mitochondrial genes [6]. Any cells that did not meet the specified threshold levels were excluded from the analysis. The thresholds applied were as follows: cells with ≥ 20,000 total genes, cells with ≤ 1000 unique genes, and cells with ≥ 10% of sequences mapping to mitochondrial genes [7,8]. First, each sample was normalized for sequencing depth. Then, we addressed batch effects using Seurat's IntegrateData function. Initially, the top 1000 variable features for each normalized sample were identified using Seurat's FindVariableFeatures function, helping to pinpoint integration features. The samples were subsequently scaled, which involved normalizing by the standard deviation for each gene and centered by subtracting the average expression for each gene. These pre-processing steps were performed prior to conducting principal component analysis (PCA) on 50 principal components (PCs). Lastly, the reciprocal PCA method was utilized to identify the integration anchors.

**Dimensionality reduction and clustering**

The UMAP algorithm was applied to the PCs to reduce the dimensionality to two for visualization purposes. The resolution parameter of Seurat's FindClusters function was set to 0.9. It was confirmed that the integrated patient dataset did not exhibit clusters that were specific to a particular batch; instead, each cluster contained cells derived from multiple samples.

**Cell type identification**

We identified cell types from the scaled and integrated expression matrix using the SingleR algorithm [9]. Cluster annotations were assigned based on the cell type with the highest SingleR score. Additionally, we manually annotated cell types by combining information from published literature and the CellMarker database (http://xteam.xbio.top/CellMarker/), relying on agreed-upon and wellknown cell markers.

**Pathway analysis of megakaryocyte subset**

We utilized the enrichplot package to perform GO pathway enrichment analysis on a megakaryocyte subset. Genes were filtered using cutoff values of p = 0.05 and q = 0.05. Next, we identified the top five most significant pathways and visualized them using a dot plot. Subsequently, we selected the important pathways that were shared with the GO pathways related to cluster 4 of dynamic plasma proteomics analysis and visualized them using a heat plot.

**Differential expression across samples and correlation analysis between genes**

We extracted gene expression levels, visualized the gene expression levels between groups with the ggpubr package (https://github.com/kassambara/ggpubr), conducted a non-parametric test between groups using the Wilcoxon test, and obtained the correlation of genes with Pearson analysis.

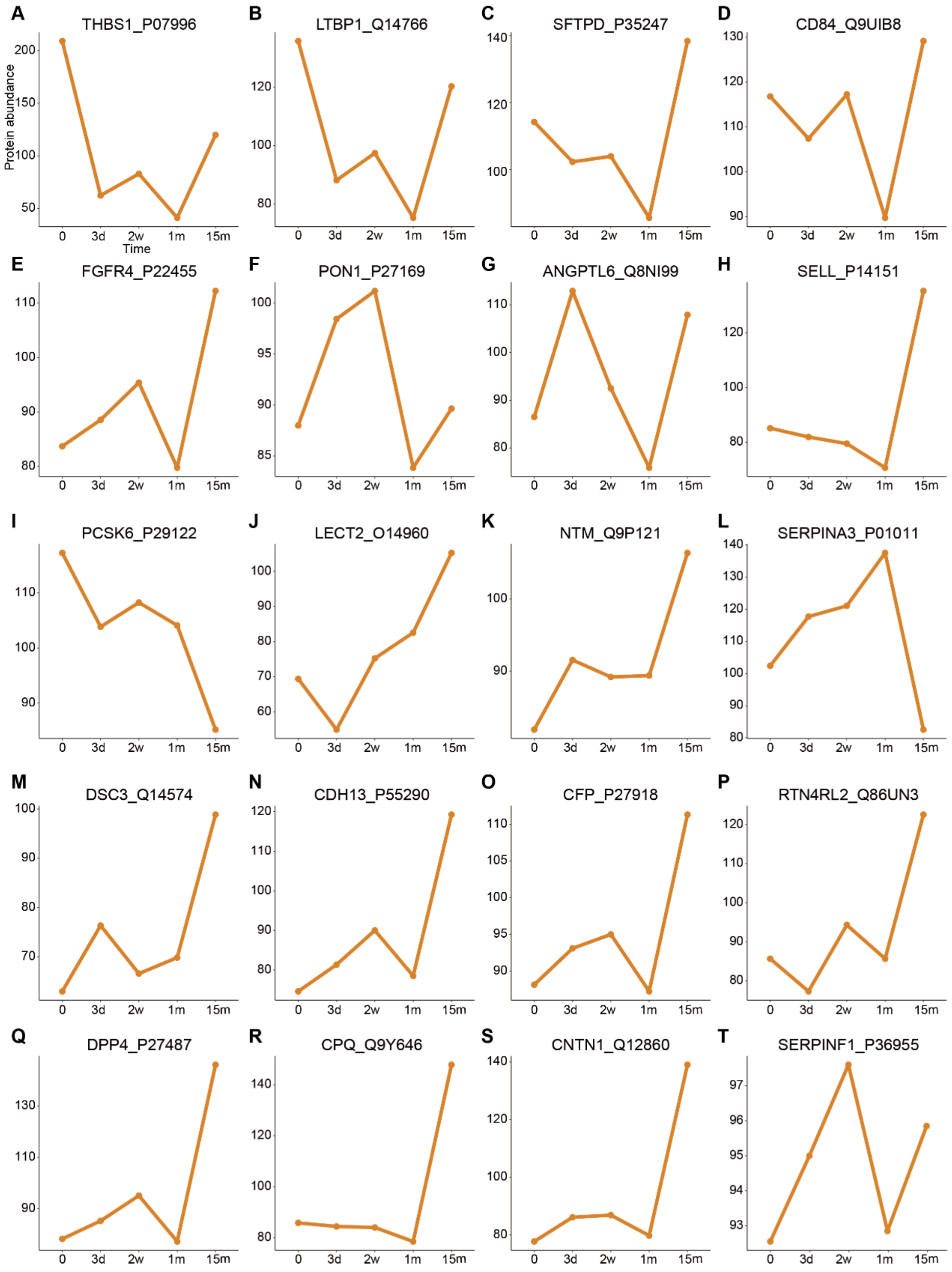


**Supplementary Fig. S1: Dynamic trends of 20 differentially expressed proteins in the plasma of CUA patient 1 treated with hAMSCs were analyzed through proteomic analysis.**

**(A)** THBS-1. **(B)** LTBP1. **(C)** SFTPD. **(D)** CD84. **(E)** FGFR4. **(F)** PON1. **(G)** ANGPTL6. **(H)** SELL. **(I)** PCSK6. **(J)** LECT2. **(K)** NTM. **(L)** SERPINA3. **(M)** DSC3. **(N)** CDH13. **(O)** CFP. **(P)** RTN4RL2. **(Q)** DPP4. **(R)** CPQ. **(S)** CNTN1. **(T)** SERPINF1.

*CUA, calcific uremic arteriolopathy; hAMSCs, human amnion-derived mesenchymal stem cells; THBS-1, thrombospondin-1; LTBP1, latent transforming growth factor beta binding protein 1; SFTPD, surfactant protein D; CD84, cluster of differentiation 84; FGFR4, fibroblast growth factor receptor 4; PON1, paraoxonase 1; ANGPTL6, angiopoietin like 6; SELL, selectin L; PCSK6, proprotein convertase subtilisin/kexin type 6; LECT2, leukocyte cell derived chemotaxin 2; NTM, neurotrimin; SERPINA3, serpin family A member 3; DSC3, desmocollin 3; CDH13, cadherin 13; CFP, complement factor properdin; RTN4RL2, reticulon 4 receptor like 2; DPP4, dipeptidyl peptidase-4; CPQ, carboxypeptidase Q; CNTN1, contactin 1; SERPINF1, serpin family F member 1.*

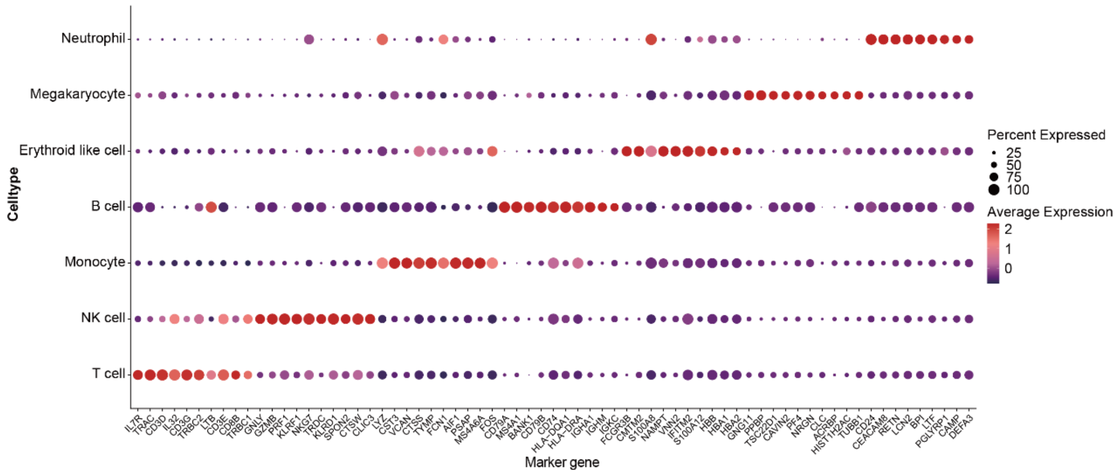


**Supplementary Fig. S2: Cell type identification in the scRNA-seq integrated datasets of PBMCs from a healthy control and CUA patient 3.**

Dot plot representing canonical PBMC markers on the x-axis. Each cluster is labeled as a specific cell type based on its expression pattern, indicated on the y-axis. The size of the dots represents the fraction of cells within the cluster that express the marker, while the color indicates the average expression level of the marker.

*scRNA-seq, single-cell RNA sequencing; CUA, calcific uremic arteriolopathy; PBMCs, peripheral blood mononuclear cells; NK cell, natural killer cell.*

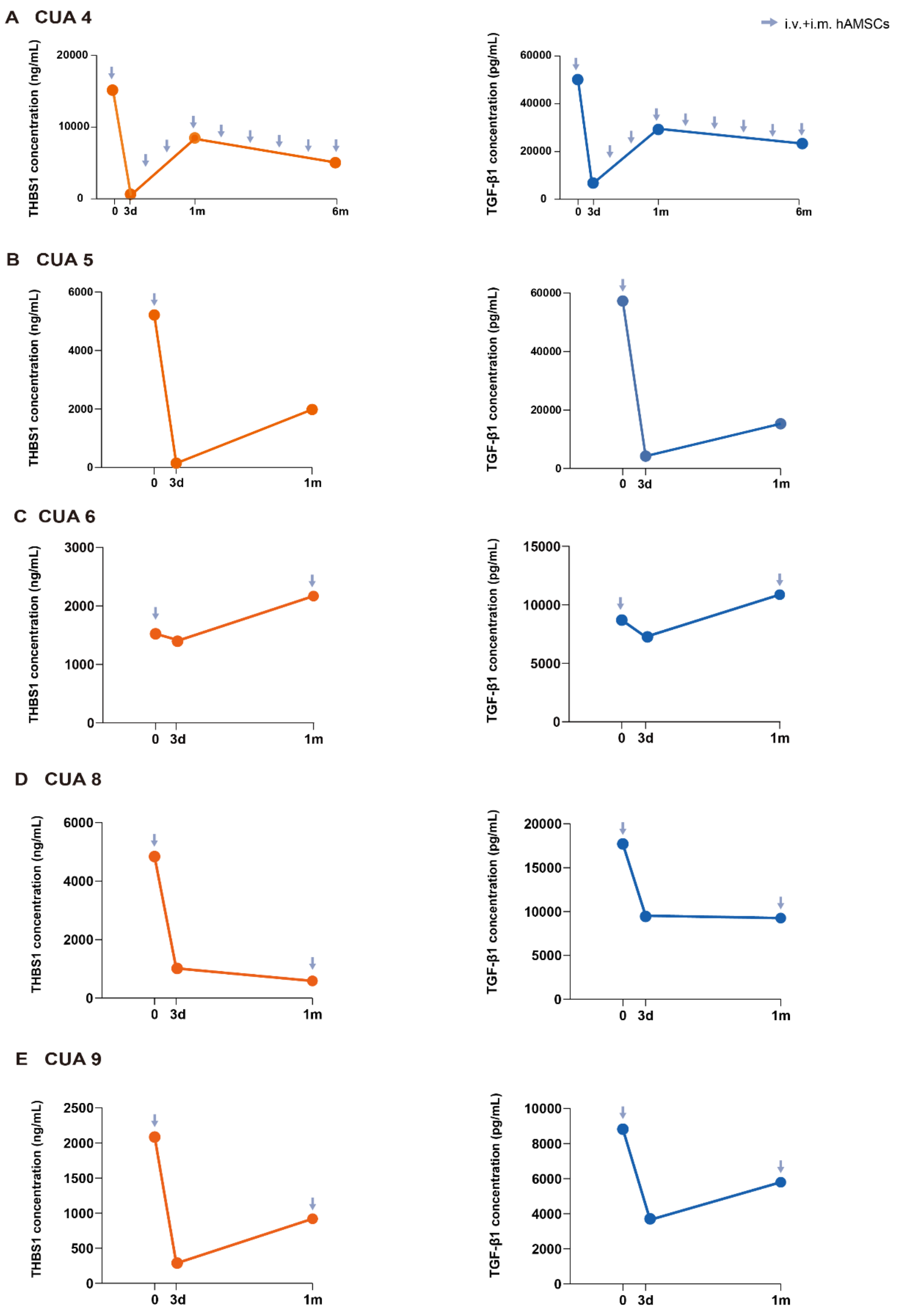


**Supplementary Fig. S3: Dynamic THBS1/TGF-β1 plasma levels of CUA patients in the validation cohort were measured by ELISA during the course of hAMSC treatment.**

**(A-E)** Plasma THBS1 (orange) and TGF-β1 (blue) levels in CUA patients 4, 5, 6, 8, and 9 were monitored throughout the duration of hAMSC treatment by ELISA.

*CUA, calcific uremic arteriolopathy; hAMSC, human amnion-derived mesenchymal stem cell; THBS1, thrombospondin 1; TGF-β1, transforming growth factor-β1; ELISA, enzyme linked immunosorbent assay.*

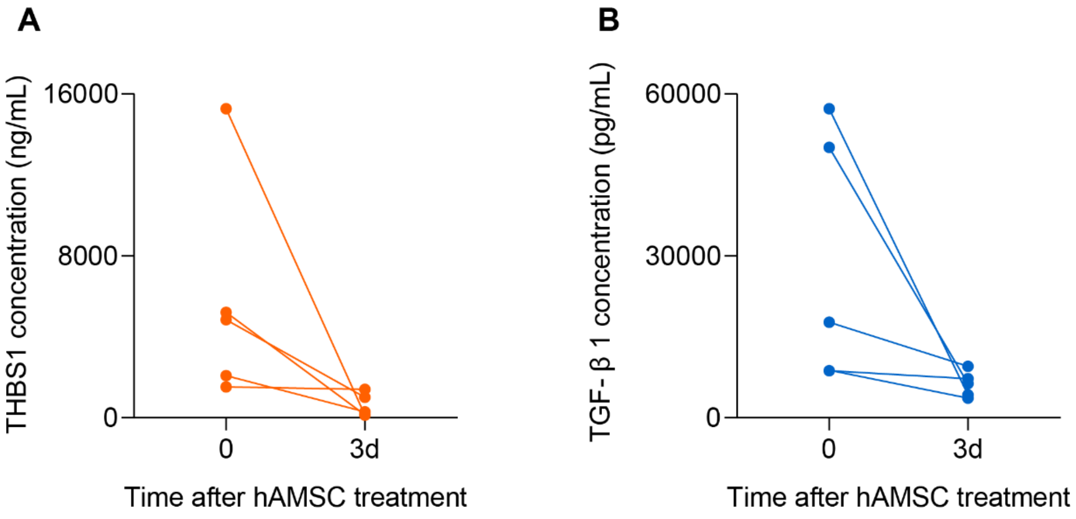


**Supplementary Fig. S4: Plasma THBS1 and TGF-β1 levels of five CUA patients were measured in the validation cohort before and after treatment with hAMSCs for 3 days.**

*CUA, calcific uremic arteriolopathy; hAMSCs, human amnion-derived mesenchymal stem cells; THBS1, thrombospondin 1; TGF-β1, transforming growth factor-β1.*

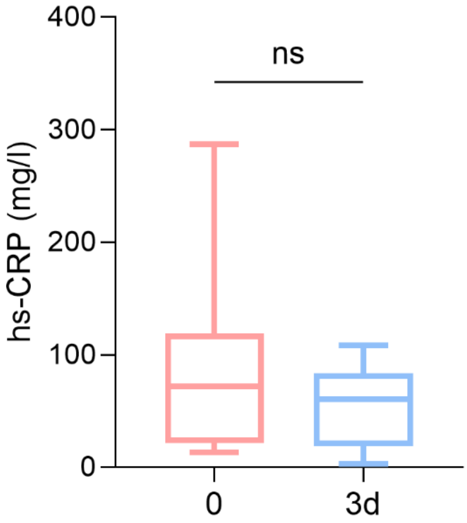


**Supplementary Fig. S5: Comparison of hs-CRP levels before and 3 days after hAMSC treatment in CUA patients.**

For the eight CUA patients treated with hAMSCs, blood hs-CRP levels were reduced 3 days after treatment compared with pre-treatment levels, but there was no statistical difference.

*hs-CRP, hypersensitive C-reactive protein.*
